# Can Vision–Language Models See the Vital Signs? Benchmarking and Fine-Tuning for Intraoperative Monitor Reading

**DOI:** 10.64898/2026.06.10.26355351

**Authors:** Gao Shaowei, Wang Pingqian, Zhao Xu, Yang Bo, Zhang Zheyi, Feng Xia, He Qiulan

## Abstract

**Background:** Vital-sign deterioration is a leading contributor to preventable perioperative death, yet manual monitor reading is intermittent, error-prone, and subject to alarm fatigue. Automating this perceptual step could enable continuous surveillance, but existing solutions depend on device-specific hardware integration or cloud-hosted vision–language models (VLMs), which raise privacy, cost, and connectivity barriers in resource-limited healthcare facilities.

**Methods:** We constructed a benchmark of 200 in-the-wild intraoperative monitor photographs (spanning multiple vendors, angles, and illumination conditions) annotated for eight vital-sign parameters: heart rate, SpO₂, ETCO₂, respiratory rate, systolic/diastolic/mean blood pressure, and temperature. We evaluated an optical character recognition (OCR)-based pipeline, nine instruction-tuned VLMs (four commercial, five open-weight ranging from ≤4B to 31B parameters) under two prompting regimes, and a compact open model (Qwen3.5-9B) adapted via low-rank fine-tuning (LoRA, 0.46% of parameters updated).

**Results:** Under a domain-aware prompt, frontier VLMs reached 0.98–0.997 exact-match accuracy zero-shot, whereas the OCR pipeline and ≤4B model scored approximately 0.20 lower, defining a 9B-class usable floor. LoRA fine-tuning Qwen3.5-9B on 80–120 images raised accuracy from 0.953 to 0.994 (statistically indistinguishable from the best commercial model) and reduced the critical-error rate fivefold (0.0313 → 0.0063). Ablations showed that performance saturated at 80 training images and rank-8 adapters.

**Conclusion:** Monitor reading is a solved perception problem for VLMs above the 9B scale. A lightweight fine-tuned open model achieves frontier accuracy while running entirely on local hardware, preserving data privacy, offline capability, and near-zero marginal cost. Residual errors stem from blood-pressure source ambiguity and are addressable with explicit disambiguation logic.

## 1. Introduction

For artificial intelligence in medicine, as for the clinician it is built to assist, the first step is to see. Before any diagnosis is weighed or intervention chosen, the relevant facts must first be perceived: the chart read, the patient examined, and above all the monitor watched — heart rate, blood pressure, oxygen saturation, respiratory rate, capnography, and temperature anchor nearly every perioperative decision. Undetected deterioration in vital signs is a leading and partly preventable contributor to perioperative morbidity and mortality, whether intraoperatively — where transient hypotension, hypoxemia, or arrhythmia can escalate within minutes — or on the general ward, where most postoperative deaths occur [1]. Detecting those changes promptly measurably improves outcomes: continuous surveillance reduced rescue events by roughly two-thirds and intensive-care transfers by nearly half relative to intermittent checks [2].

Yet human reading is the weakest link precisely where it matters most. Heavy workload and the documentation burden that drives clinician burnout erode sustained attention [3]. An estimated 85–99% of monitor alarms are false or clinically non-actionable, and the resulting alarm fatigue desensitizes staff so that genuine, actionable events are delayed or missed [4]. Because manual charting is intermittent, the slow physiological drift that heralds deterioration goes unrecorded between checks. Automating the act of reading the monitor is therefore a question of patient safety, not of convenience.

In addition to safety, data accessibility is itself a clinical need: vital signs locked inside a device cannot reach the electronic health record (EHR) or the early-warning scores that depend on them. In resource-limited healthcare facilities, legacy monitors were never designed to export data. Direct hardware or vendor software development kit (SDK) integration is costly, device-specific, and rarely feasible to retrofit across a heterogeneous fleet [5]. Sending screen photographs to a commercial vision–language model (VLM) is technically capable but often clinically inadmissible: it raises data-sovereignty and privacy concerns and incurs a recurring per-query cost. A practical solution must therefore read in-the-wild photographs accurately while running on-premises, offline, and at low marginal cost.

We test whether this perceptual step can be reliably automated under those constraints through three hypotheses. First, that a contemporary vision–language model can read an intraoperative monitor from an ordinary photograph with accuracy sufficient to be clinically useful — and that the model capacity the task requires can be empirically delineated rather than assumed. Second, that in settings requiring offline operation and data privacy, a compact open-weight model can be adapted through lightweight fine-tuning from a modest amount of labelled data. Third, that the resulting readings can be rendered clinically trustworthy, judged not by average accuracy but by whether they avoid the specific errors that would alter a clinical decision. To test these hypotheses, we construct a controlled benchmark from real monitor photographs and evaluate a hierarchy of methods spanning a classical OCR pipeline to small, open, and frontier vision–language models, then adapt a compact open model with low-rank fine-tuning [6].

## 2. Materials and Methods

We study the digitization of vital signs displayed on bedside patient monitors as a structured visual-information-extraction problem: given a single photograph of a monitor screen, a method must output the numerical value of each of eight vital-sign fields, or report that the field is absent.

### 2.1 Dataset and annotation

#### Image acquisition

The study was approved by the Clinical Research and Laboratory Animal Ethics Committee of the First Affiliated Hospital of Sun Yat-sen University (approval no. [2025]129). The dataset comprises 200 photographs of bedside patient-monitor screens, acquired with handheld smartphones during surgical procedures. Each photograph is a single-moment snapshot of the monitor display; no patient-identifiable information (name, medical record number, or other protected health information) appears in any image. The photographs reflect routine intraoperative conditions rather than a controlled capture setup: they vary in viewing angle, distance, illumination, screen glare, and reflection, and they span multiple monitor vendors, models, and on-screen layouts. The image set is heterogeneous in both content and file format— as an incidental but realistic property, 98 of the 200 files carry a .png extension while actually being JPEG-encoded, and 5 files exceed 5 MB (up to 25 MB) — so any practical pipeline must be robust to in-the-wild data. To handle this heterogeneity, our pipeline detects actual image format from file-header magic bytes rather than from the file extension, and downscales images exceeding 1,536 px on their longest side to stay within model input limits. The distribution of monitor brands and models is summarized in **Table 1**; the cohort is dominated by Mindray devices (notably the BeneView T8 and BeneVision N-series), with a minority of unidentifiable units.

**Table 1.** Distribution of display brands and models, along with the number of images provided.

| brand | model | n_images |
| --- | --- | --- |
| Mindray | BeneView T8 | 86 |
| Mindray | BeneVision N-series | 70 |
| Mindray | Mindray (unspecified) | 22 |
| Unknown | Unknown | 19 |
| Mindray | BeneView (other) | 2 |
| Mindray | BeneVision | 1 |

#### Target fields

Each image is annotated for eight vital-sign parameters: heart rate (HR, bpm), peripheral oxygen saturation (SpO₂, %), end-tidal carbon dioxide (ETCO₂, mmHg), respiratory rate (RR, breaths/min), systolic blood pressure (SBP, mmHg), diastolic blood pressure (DBP, mmHg), mean arterial pressure (MAP, mmHg), and body temperature (TEMP, °C). Not every parameter is shown on every monitor; a field that is not displayed is annotated as *absent* (null) rather than guessed, and a method is credited for that field only if it also returns null.

This yields 200 × 8 = 1,600 parameter instances, a meaningful fraction of which are legitimately absent (most commonly ETCO₂, MAP, and TEMP), so that correctly abstaining is itself part of the task.

#### Reference standard

Ground-truth values were established through a structured consensus process: two researchers independently read each monitor screen and recorded the values in a standardized table; discrepancies were adjudicated by a third researcher to produce the final reference.

### 2.2 Data partition

We partition the 200 images at the image level into training, validation, and test subsets in a 6:2:2 ratio (120 / 40 / 40). To keep the three subsets comparable in difficulty, the split is stratified by *information density*, defined as the number of non-absent parameters on an image, so that screens displaying many simultaneous parameters (typically more advanced monitors) are represented proportionally in each subset; the partition uses a fixed random seed (42) for reproducibility. The validation subset is used exclusively for model and checkpoint selection and for early stopping during fine-tuning; the test subset is held out and scored exactly once, for the final results reported in the Results section. The exact image-to-subset assignment is released with the code.

### 2.3 Models and prompting strategies

We compare three families of models, all producing the same eight-key JSON output.

#### OCR pipeline

As a lightweight, fully open baseline we build a two-stage pipeline. In preliminary experiments we tested three OCR engines: Tesseract 5, a conventional OCR system based on long short-term memory–recurrent neural network (LSTM-RNN) line recognition trained predominantly on printed and handwritten document fonts, produced near-zero output on monitor images — its models have no representation of the color-coded seven-segment and proprietary LED-style digits used by bedside monitors, and its line-based segmentation fails on the multi-region, non-linear layouts of monitor screens.

PaddleOCR-VL-1.5, a 0.9B-parameter generative vision–language model (NaViT encoder + ERNIE-4.5-0.3B decoder) [7], suffered from repetition degeneration: rather than recognizing screen content, it produced infinite loops of a few tokens until the output budget was exhausted — a known failure mode of small autoregressive models on out-of-distribution visual inputs. We therefore adopted PaddleOCR PP-OCRv4 [7], which uses a convolutional neural network (CNN)-based detector (DBNet) and a hybrid CNN–Transformer recognizer (SVTR-LCNet) to localize and recognize text fragments with bounding-box coordinates. These unstructured fragments are then mapped to the eight-field JSON by an open large language model (LLM) used as a post-processor (Qwen3.5-9B, non-reasoning mode, temperature 0). The post-processor receives only the recognized text and spatial coordinates; it never sees the original image, which isolates the contribution of optical recognition from that of structured parsing. We evaluate this pipeline under both prompting regimes described below (applied to the post-processing stage).

#### Vision–language models

We evaluate nine instruction-tuned vision–language models spanning a wide capability and size range: four proprietary frontier models accessed through hosted APIs (GPT-5.5, Claude-Opus-4.8, Gemini-3.1-Pro, Gemini-3.5-Flash) and five open-weight models (Qwen3.6-27B, Qwen3.5-9B [8], GLM-5V-Turbo [9], Gemma-4-31B, and the compact ≤4B model Gemma-4-E4B [10]). All models are queried through OpenAI-compatible endpoints at temperature 0 for deterministic, comparable outputs; the per-request token budget is raised for the reasoning-style models (the Gemini models and the Qwen3.5/3.6 series, which emit explicit reasoning traces) to avoid truncation of their final answer. To accommodate the heterogeneous inputs noted in the Dataset section, images are dispatched with the content type inferred from their file-header bytes (not their file extension) and are downscaled to a longest side of 1,536 px when they exceed provider size limits.

#### Prompting regimes

Each method is run under two prompts. The *basic* prompt states only the task and the required JSON keys. The *advanced* prompt is domain-aware: it additionally specifies, for each field, its on-screen label conventions, unit, and the disambiguation rules that monitor screens require — for example, that SBP and DBP are the upper and lower numbers of a blood-pressure reading and that temperature may appear as T, T1, or T2 and excludes alarm-limit values. Comparing the two regimes isolates how much of each method’s performance comes from task framing versus from the model’s intrinsic capability. The verbatim prompts are provided in Appendix A.

### 2.4 Evaluation protocol and metrics

All methods emit, for each image, a JSON object with the eight parameter keys; we parse this output robustly and normalize each value to a number or null before scoring. Unless otherwise stated, scoring is performed at the level of the parameter instance (1,600 instances overall; 320 on the held-out test set). All metric definitions and statistical procedures are implemented in a single shared scoring module used uniformly across every method, so that all reported numbers share one scoring convention.

#### Exact match

Our primary metric is exact-match accuracy (EM): a parameter instance is correct when the predicted value equals the reference value, with null required to match null. Because EM is a proportion measured on a finite sample, we accompany every EM figure with a 95% Wilson score confidence interval [11, 12], which is better calibrated than the normal approximation in the high-accuracy regime relevant here.

#### Clinical metrics

Exact match treats a one-unit error and a clinically dangerous error identically. To assess clinical validity we therefore add three complementary analyses over the numeric parameters. (i) *Tolerance accuracy* credits a prediction as correct when it falls within a clinically negligible margin of the reference value; we use HR ±3 bpm, SpO₂ ±2%, SBP/DBP/MAP ±5 mmHg, TEMP ±0.3 °C, ETCO₂ ±3 mmHg, and RR ±2 breaths/min. (ii) The *critical-error rate* is the fraction of parameter instances whose error is large enough to plausibly alter a clinical judgement— operationalized as an absolute deviation exceeding a per-parameter critical threshold (HR 15 bpm, SpO₂ 4%, ETCO₂ 8 mmHg, RR 6 breaths/min, SBP 15 mmHg, DBP 10 mmHg, MAP 12 mmHg, TEMP 1.0 °C), or a missed detection (a displayed value read as null) or a false detection (an absent field assigned a value). (iii) *Bland–Altman analysis* [13], the standard tool for method-comparison in measurement, characterizes the bias and 95% limits of agreement between predicted and reference values; we report it for HR, SBP, and SpO₂ as representative parameters.

#### Statistical comparison

Pairwise comparisons between methods on the same images use the exact-binomial McNemar test [14], which conditions on the images where the two methods disagree and is appropriate for paired, correlated predictions. For inter-method comparisons (the model comparison subsection of the Results) the test is two-sided. For the prompt-effect comparison (the prompt-engineering subsection of the Results), where the domain-aware advanced prompt is expected a priori to perform at least as well as the minimal basic prompt, we use a one-sided variant (H₁: advanced ≥ basic): under the null hypothesis the discordant pairs follow a Binomial(*n*, 0.5) distribution, and the *p*-value is the upper-tail probability of observing at least as many basic-wrong → advanced-right switches as actually occurred.

### 2.5 Lightweight LoRA fine-tuning

#### Recipe

To obtain a model that is accurate yet deployable under the offline, privacy-constrained conditions of non-integrated wards, we fine-tune the compact open-weight Qwen3.5-9B with Low-Rank Adaptation (LoRA) [6].

Adapters of rank 16 (α = 32, dropout 0.05) are applied to all linear projections of the language model while the vision encoder and the vision–language aligner are kept frozen; this updates 43.3 M parameters, 0.46% of the model. Fine-tuning is performed with the MS-SWIFT framework [15] for 5 epochs at learning rate 1×10⁻⁴ (cosine schedule, 5% warmup, weight decay 0.1), effective batch size 8 (per-device batch 2 with gradient accumulation 4), in bfloat16 with scaled-dot-product attention and a 4,096-token context; the random seed is fixed at 42. We use only the 120 training images defined in the Data Partition section; the advanced prompt serves as the instruction, and the model is trained and queried in its direct-answer (non-reasoning) mode, which both matches the JSON-only training targets and is the configuration intended for deployment.

#### Model selection and evaluation

Checkpoints are evaluated on the validation subset every 15 optimization steps, and the checkpoint with the lowest validation loss is selected for testing— deliberately *not* the checkpoint with the lowest training loss, since training loss collapses early (see the fine-tuning subsection of the Results). Here the validation loss is minimized at step 30 (the second of five epochs), after which it rises again, so checkpoint-30 is selected; the selected checkpoint is then run once on the held-out test set at temperature 0. As a factual record of cost, the full 75-step run took roughly 26 minutes on a single NVIDIA RTX Pro 5000 (48 GB) GPU rented through the Vast.ai cloud platform, with a measured peak memory of 28.5 GB.

#### Ablations

Two ablations probe the recipe’s data and capacity requirements, both conducted on the same Qwen3.5-9B backbone and under the identical protocol as the main recipe. The data-efficiency ablation re-runs fine-tuning on nested training subsets of 0 (zero-shot base), 40, 80, and 120 images, with all other settings fixed, to locate the point of diminishing returns. The rank ablation varies the LoRA rank over 8, 16, 32, and 64 (with α = 2 × rank) at the full 120-image training size. In every ablation run the checkpoint is selected on validation loss and the test set is scored once, exactly as for the main recipe. When no ablation configuration yields a clear improvement over the default settings (120 images, rank 16), the result from the default configuration is used for all subsequent evaluations.

## 3. Results

### 3.1 Performance landscape across models and the role of prompt engineering

We evaluated each model zero-shot on all 200 images under two prompting conditions: a minimal *basic* prompt and a domain-aware *advanced* prompt that specifies vital-sign parameter names, expected units, and output format *(detailed in Section 2)*. Results appear in **Fig. 2A**, with a per-parameter breakdown in **Fig. 2B** and exact-match accuracies with 95% Wilson confidence intervals in **Table S1**. One-sided McNemar tests for the prompt effect (H₁: advanced ≥ basic) are reported in **Table S2**.

Under the advanced prompt, three performance tiers emerge (**Fig. 2A**). Four commercial models and one open-weight model form the top tier, all exceeding 0.97 exact-match accuracy: Claude-Opus-4.8 (0.984), GPT-5.5 and Gemini-3.1-Pro (0.981 each), Gemma-4-31B (0.976), and Gemini-3.5-Flash (0.974). Notably, Gemma-4-31B — an open-weight model — sits comfortably within the commercial range, showing that an open-weight model already matches the best commercial models on this task.

**Figure 1.**
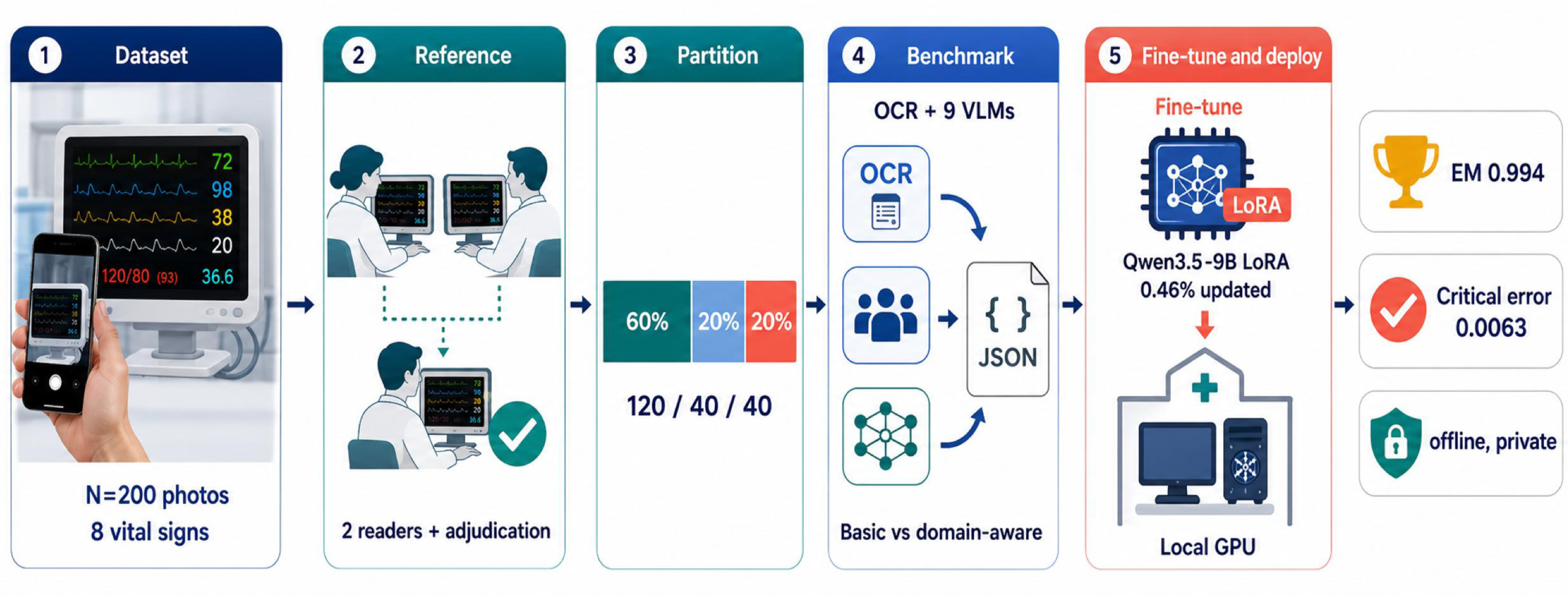
Study workflow. (1) Two hundred intraoperative monitor photographs were collected from routine surgical procedures. (2) Ground-truth values for eight vital-sign parameters were established by dual-reader annotation with third-reader adjudication. (3) Images were split 6:2:2 into training, validation, and test subsets, stratified by information density. (4) An OCR-based pipeline and nine vision–language models were benchmarked under basic and domain-aware prompts. (5) Qwen3.5-9B was fine-tuned with LoRA (0.46% of parameters) and deployed on a local GPU, achieving 0.994 exact-match accuracy and a critical-error rate of 0.0063.

**Figure 2.**
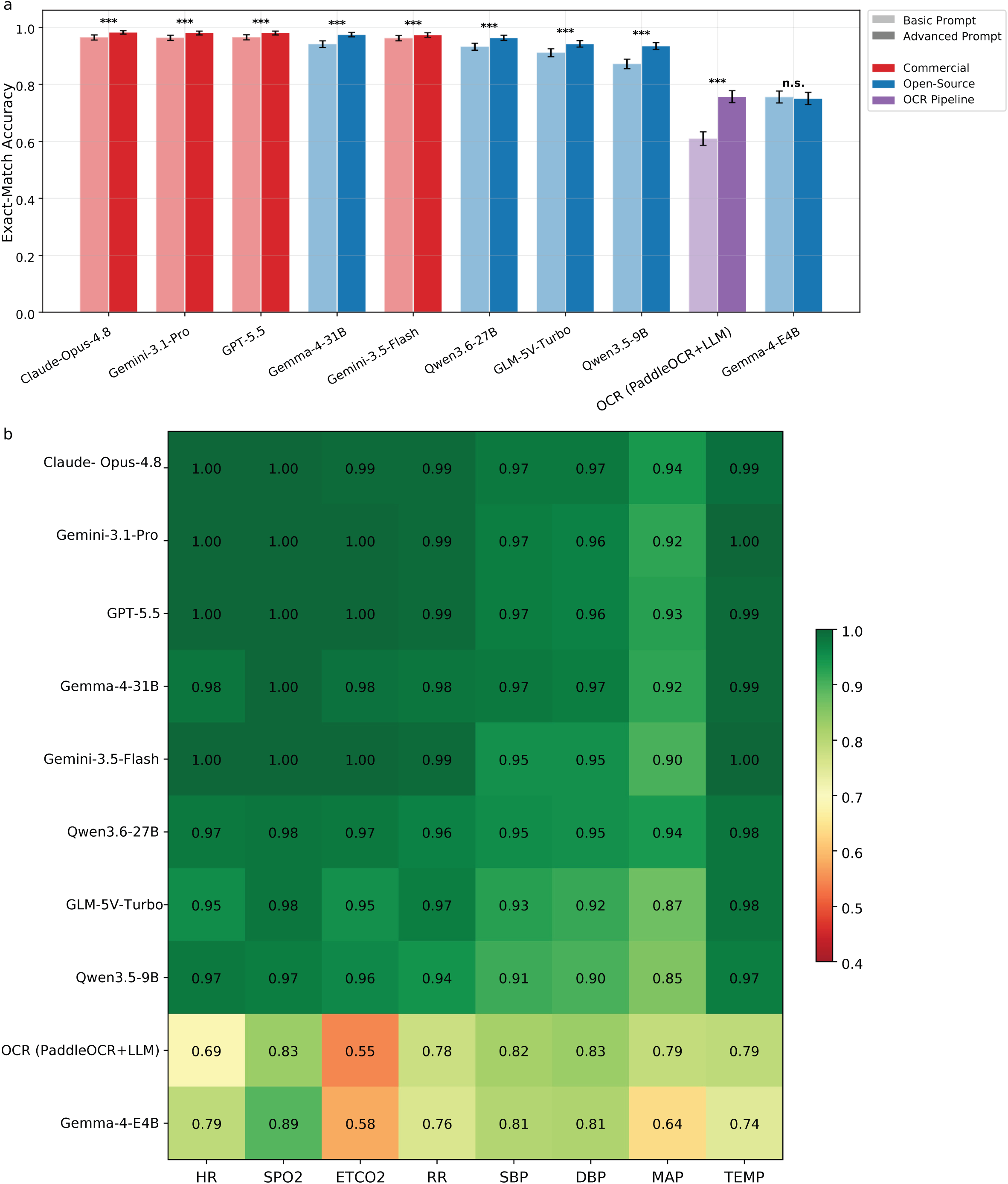
Zero-shot performance landscape. (**a**) Exact-match accuracy of all evaluated methods on the full 200-image dataset under basic (light bars) and advanced (dark bars) prompts. Methods are colored by category: commercial API (red), open-weight VLM (blue), and OCR pipeline (purple). Significance stars indicate one-sided McNemar tests for the prompt effect (H₁: advanced ≥ basic): *** *p* < 0.001, n.s. *p* ≥ 0.05. Error bars denote 95% Wilson confidence intervals. (**b**) Per-parameter exact-match heatmap under the advanced prompt. Rows are models (ordered by overall accuracy); columns are the eight vital-sign parameters. Color scale ranges from 0.4 (red) to 1.0 (green).

A second tier of mid-sized open models follows at 0.94–0.96: Qwen3.6-27B (0.964), GLM-5V-Turbo (0.943), and Qwen3.5-9B (0.936). These remain competent but fall measurably below the leaders.

Below this tier, accuracy drops sharply rather than tapering off. The LLM-based OCR pipeline (0.746) and the compact Gemma-4-E4B (0.751) lie roughly 20 points behind the mid-tier VLMs, well beyond any clinically acceptable margin.

This OCR pipeline already relies on the same 9B language model (Qwen3.5-9B) for structured extraction, yet its bottleneck lies upstream: PP-OCRv4’s character-level recognition produces incomplete or erroneous fragments from the multi-colored, non-standard fonts of monitor screens, and no amount of downstream LLM reasoning can recover information that was never extracted. Conventional OCR engines fare even worse — Tesseract 5 produces near-zero output because its LSTM recognizer, trained on document fonts, has no representation of seven-segment LED digits, and the generative PaddleOCR-VL-1.5 (0.9B) collapses into degenerate repetition rather than reading the screen. This cliff, rather than the narrow spread among leaders, defines the practical structure of Fig. 2A: the usable floor for this task lies at the 9B-class VLM, and models below it are not marginally but categorically worse.

Prompting interacts with model capability in a revealing pattern (**Table S2**). Even for top-tier models, where the absolute gain is modest, the one-sided McNemar test confirms that the improvement is highly significant: GPT-5.5 (0.966→0.981, *p* = 5.96 × 10⁻⁸), Claude-Opus (0.966→0.984, *p* = 6.54 × 10⁻⁸). In both cases 24–31 parameter instances switch from wrong to right with zero to two moving in the opposite direction — a near-perfectly asymmetric shift despite the small absolute magnitude. Mid-tier models benefit more and with even greater significance (Qwen3.5-9B: 0.873→0.936, *p* = 8.03 × 10⁻²¹), and the effect is most pronounced in the LLM-based OCR pipeline (0.677→0.746, p = 1.83 × 10⁻¹²): the prompt supplies the domain context that pixel-level processing alone cannot recover. The compact Gemma-4-E4B, by contrast, shows no benefit from prompting (basic 0.756 vs. advanced 0.751, *p* = 0.69), confirming that its bottleneck lies in visual recognition capacity before domain knowledge even matters.

Prompt engineering thus narrows but cannot close the capability gap: it improves weaker models but does not lift them onto the top-tier plateau. The rest of this paper fixes the advanced prompt and asks what else is needed beyond prompting.

### 3.2 Fine-tuned Qwen3.5-9B reaches frontier-level accuracy

To test whether lightweight fine-tuning can close the gap that prompting alone cannot, we applied LoRA to Qwen3.5-9B, a mid-tier 9B model. Training on 120 images raises its exact-match accuracy from 0.953 (zero-shot) to **0.994** (95% Wilson CI 0.978–0.998), a **+4.1-point** gain achieved by updating only 43.3 M parameters (0.46% of the model). The best checkpoint is selected by validation loss at epoch 2, step 30 (**Fig. 3A**).

**Figure 3.**
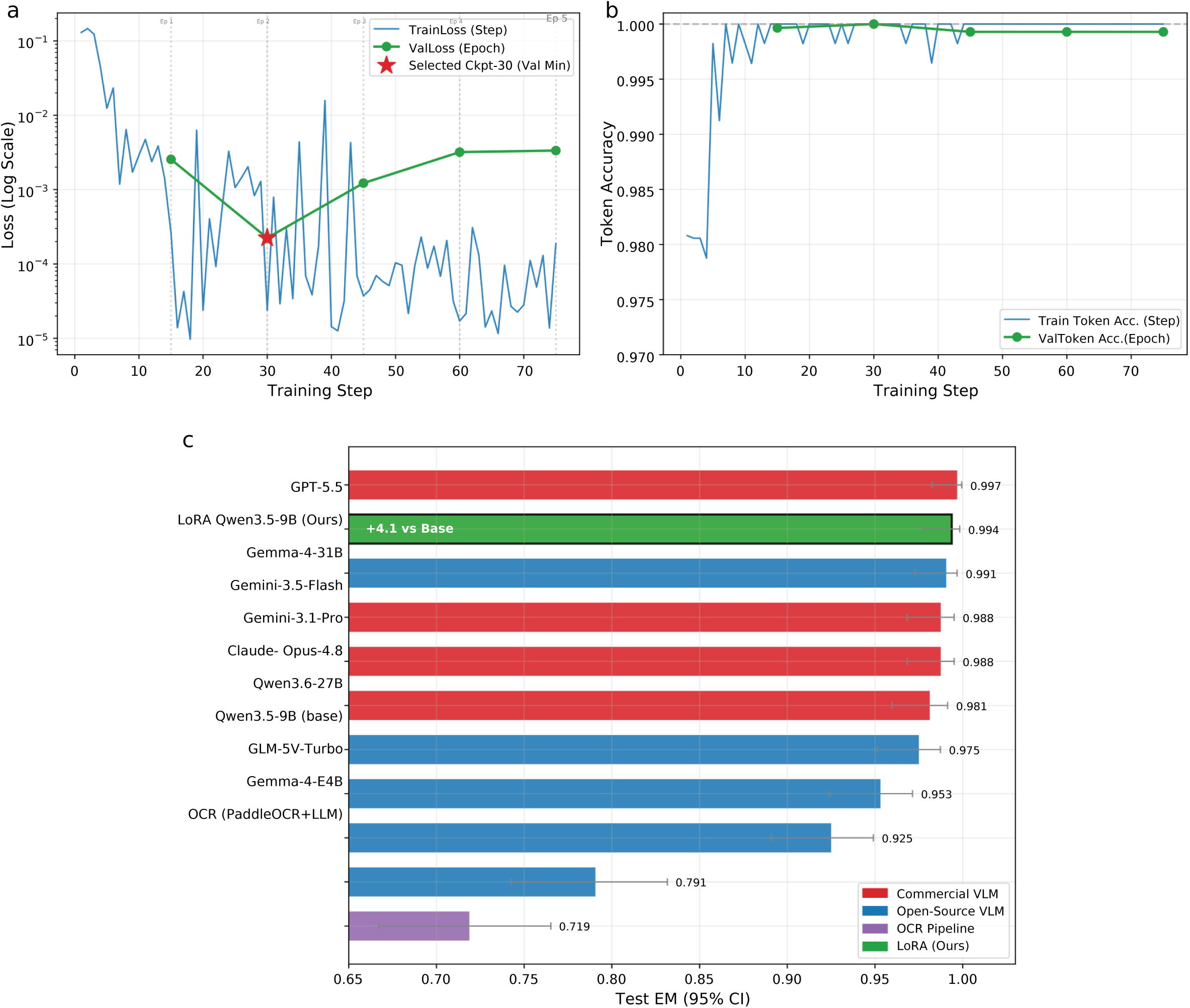
LoRA fine-tuning of Qwen3.5-9B. (**a**) Training dynamics. Left: training loss (per step, blue) and validation loss (per epoch, green) on a log scale; the red star marks the selected checkpoint (step 30, epoch 2, validation-loss minimum). Right: token accuracy for training (blue) and validation (green), showing saturation near 100% consistent with overfitting beyond epoch 2. (**b**) Method comparison on the held-out test set (40 images, advanced prompt). Horizontal bars show exact-match accuracy with 95% Wilson confidence intervals. The fine-tuned model (green, “Ours”) achieves 0.994, ranking second overall and statistically indistinguishable from GPT-5.5 (0.997; McNemar *p* = 1.0).

On the held-out test set (**Fig. 3B**), the fine-tuned 9B model ranks **second overall**, surpassing Claude-Opus-4.8 and both Gemini models and trailing only GPT-5.5 by 0.3 points — one parameter instance out of 316 valid instances (40 test images × 8 parameters, minus 4 missing ground-truth values), a difference that is not statistically significant (two-sided McNemar exact test, *p* = 1.0).

A 0.46% adapter trained on 120 images and deployable on a single local GPU thus closes the accuracy gap to the best commercial model to clinical irrelevance.

### 3.3 Ablation studies: training data size and LoRA rank

To verify that our choice of training set size (120 images) and LoRA rank (16) is reasonable rather than arbitrary, we ran two ablations on the same Qwen3.5-9B backbone (**Fig. 4**).

**Figure 4.**
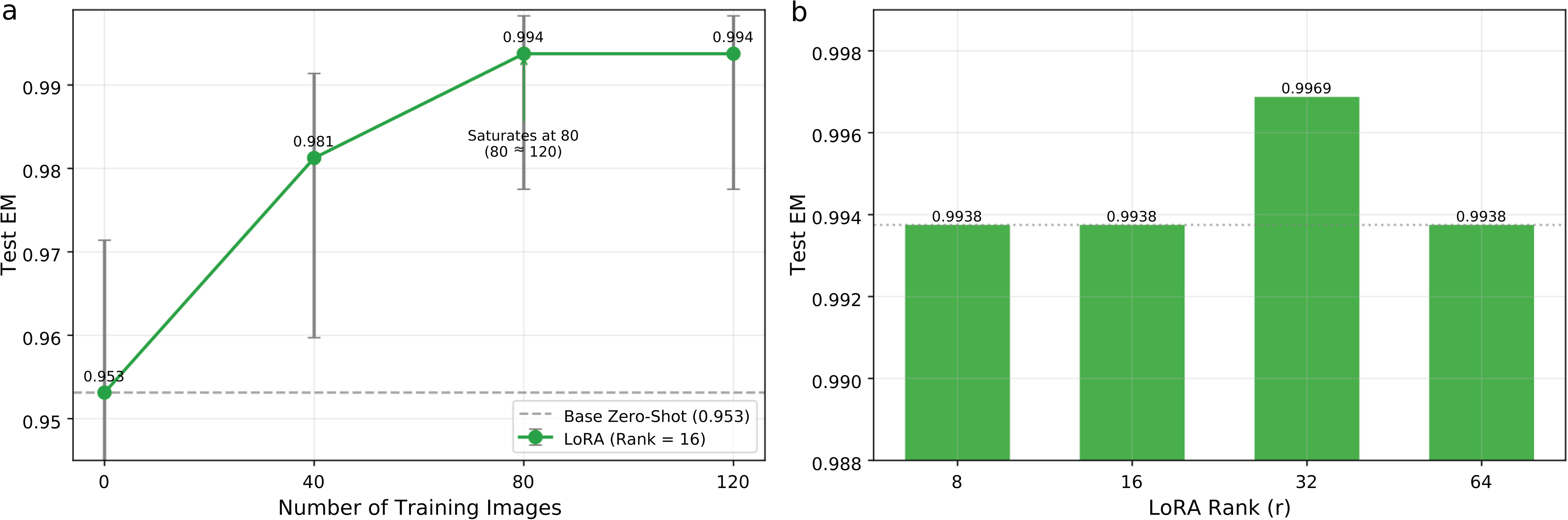
Ablation studies. (**a**) Data-efficiency ablation. Test exact-match accuracy as a function of training set size (0, 40, 80, 120 images) at fixed LoRA rank 16. Performance saturates at 80 images (0.994), with no further gain from the additional 40. The dashed line indicates the zero-shot baseline (0.953). Error bars denote 95% Wilson confidence intervals. (**b**) Rank ablation. Test exact-match accuracy at LoRA ranks 8, 16, 32, and 64 with the full 120-image training set. Accuracy is stable across all ranks (0.994–0.997), confirming that rank 8 already suffices.

Increasing the training set from 0 → 40 → 80 → 120 images raises test accuracy from 0.953 → 0.981 → 0.994 → 0.994. Performance climbs sharply through 80 images and then **saturates** — the last 40 add nothing. Around 80 labeled screens is enough, which matters when every annotated image costs clinician time.

The rank ablation tells a similar story: at ranks 8, 16, and 64 accuracy is identical (0.994), and rank 32 differs by a single instance (0.997 vs. 0.994). **Rank 8 already suffices**; a larger adapter brings no benefit.

Both ablations confirm that our default settings (120 images, rank 16) already operate in the saturation region, and reducing either to 80 images or rank 8 would yield the same accuracy.

### 3.4 Clinical agreement and error analysis

Exact match treats a one-digit-off reading the same as a hundred-point miss. To check whether the remaining errors are clinically significant, we examine the leading methods with agreement and safety metrics (**Figs. 5 and 6**).

**Figure 5.**
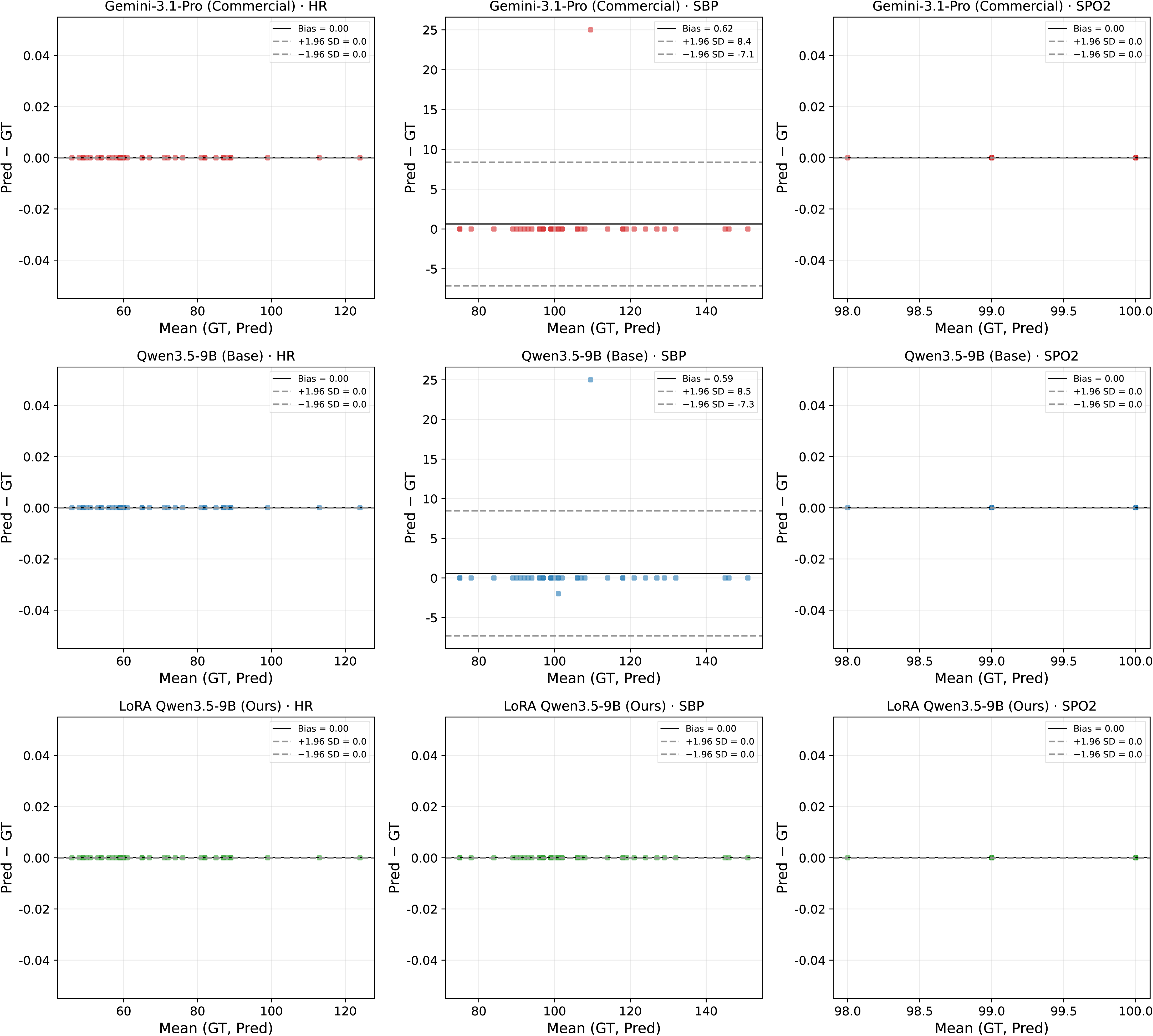
Bland–Altman agreement plots. Each panel shows the difference between predicted and reference values (y-axis) against their mean (x-axis) for a representative commercial model (Gemini-3.1-Pro, top row), the base Qwen3.5-9B (middle row), and the LoRA fine-tuned model (bottom row), across HR (left), SBP (center), and SpO₂ (right). Solid lines indicate mean bias; dashed lines indicate ±1.96 SD limits of agreement. The fine-tuned model shows near-zero bias and the tightest limits of agreement; residual spread in SBP reflects invasive-vs-non-invasive blood-pressure source confusion.

**Figure 6.**
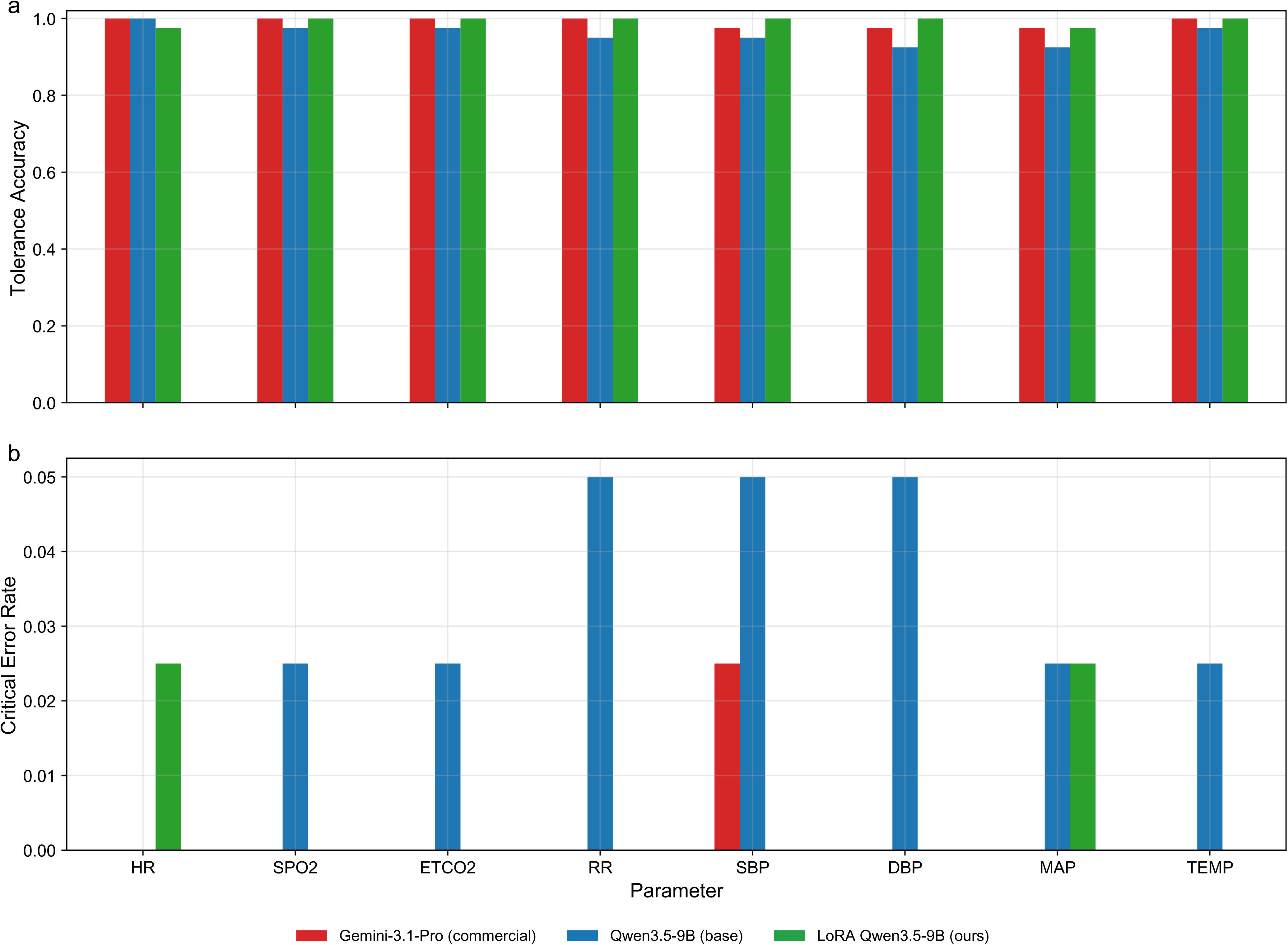
Clinical safety metrics on the held-out test set. (**a**) Tolerance accuracy by parameter for Gemini-3.1-Pro (red), base Qwen3.5-9B (blue), and fine-tuned Qwen3.5-9B (green). Clinically negligible tolerance margins: HR ±3 bpm, SpO₂ ±2%, ETCO₂ ±3 mmHg, RR ±2 breaths/min, SBP/DBP/MAP ±5 mmHg, TEMP ±0.3 °C. (**b**) Critical-error rate by parameter for the same three methods. Critical thresholds: HR 15 bpm, SpO₂ 4%, ETCO₂ 8 mmHg, RR 6 breaths/min, SBP 15 mmHg, DBP 10 mmHg, MAP 12 mmHg, TEMP 1.0 °C. Fine-tuning reduces the overall critical-error rate from 0.0313 to 0.0063, eliminating critical errors for SBP, DBP, RR, ETCO₂, SpO₂, and TEMP.

Bland–Altman plots (**Fig. 5**) are shown for HR, SBP, and SpO₂— three parameters spanning cardiac, hemodynamic, and oxygenation domains and covering the widest error range in our data. The fine-tuned model has the tightest limits of agreement among all compared methods, with nearly every point on the zero line. The residual spread concentrates in SBP, attributable to occasional invasive-vs-non-invasive blood-pressure confusion rather than digit misreading (Section 4).

Two safety metrics in **Fig. 6** address error severity directly. Tolerance accuracy (**Fig. 6A**), which credits predictions within a clinically negligible margin, shows that the fine-tuned model’s few remaining EM misses are near-misses; its mean absolute error across test parameters is effectively zero.

The critical-error rate (**Fig. 6B**) falls from **0.0313 in the base model to 0.0063 after fine-tuning** — a fivefold reduction. Per-parameter, the base model’s critical errors cluster in blood pressure (SBP, DBP, MAP) and respiratory rate; the fine-tuned model eliminates critical errors for SBP, DBP, RR, ETCO₂, SpO₂, and TEMP, leaving only two residual single-instance errors (one HR false positive, one MAP miss).

Fine-tuning thus removes the specific failure modes most likely to alter a clinical decision.

## 4. Discussion

This study yields three findings. Under a domain-aware prompt, both frontier commercial models and the strongest open model reach 0.98–0.997 exact-match accuracy zero-shot; monitor reading is a solved perception problem at this model scale. What stands out in the results is the sharp drop below the leaders: the OCR pipeline and the ≤4B model score roughly 0.20 lower, outside any plausible clinical tolerance. A 9B-class VLM marks the usable floor for this task; anything smaller fails outright.

Lightweight fine-tuning of a compact open model closes the gap to the frontier. A LoRA adapter updating 0.46% of the parameters of Qwen3.5-9B, trained on about 80 images, raises its accuracy from 0.953 to 0.994 (second among all evaluated systems, statistically indistinguishable from the best commercial model). Both the data-efficiency and rank ablations saturate early, confirming that a small amount of task-specific adaptation drives the gain rather than data volume or adapter capacity. Fine-tuning also delivers a clinical payoff: the critical-error rate falls roughly fivefold, and the blood-pressure and respiratory-rate failure modes are eliminated.

We carefully analyzed the recognition errors across all evaluated models and identified three recurring failure modes that account for both the capability cliff and the residual risk of the fine-tuned model.

i. *ETCO₂↔RR confusion.* Capnography panels place the end-tidal CO₂ value and the respiratory rate side by side. Weaker models routinely swap them, consistent with ETCO₂ and RR being the weakest cells of the per-parameter heatmap (Fig. 2B) and the dominant errors of the OCR and ≤4B baselines.
ii. *Invasive versus non-invasive blood pressure.* Monitors may display both an arterial-line and a cuff pressure. Selecting the wrong one produces a numerically plausible but large error, and is the principal source of the residual SBP spread in the Bland–Altman plot (Fig. 5). This mode is also the main residual risk that survives fine-tuning.
iii. *Glare and shadow occlusion.* Handheld capture under operating-room lighting occasionally washes out or partially occludes a digit. This mode affects even the strongest models and cannot be addressed by prompting or fine-tuning, because the information is physically absent from the image.

The first two are semantic disambiguation errors that domain-aware prompting and fine-tuning largely resolve. The third is a data-acquisition limit and sets a ceiling on what any method can achieve with in-the-wild photographs.

All prior work on extracting readings from medical device displays has followed a multi-stage detect-then-OCR paradigm. Early systems built bespoke pipelines for single-value point-of-care devices (glucometers, blood-pressure cuffs), reaching roughly 90–100% digit accuracy within a target device class [16, 17, 18]. For intensive care unit (ICU) equipment specifically, Jeon et al. (2023) built an embedded CNN-based digit recognition system (ROMI) deployed on NVIDIA Jetson platforms for reading LCD screens of standalone ICU devices such as ventilators and infusion pumps (0.989 mean average precision [mAP]) [19], and Froese et al. (2021) demonstrated Tesseract-OCR-based data extraction from a single intravenous pump model as a proof of concept, without reporting systematic accuracy metrics [20]. The most directly comparable study is that of Chau et al. (2025), who constructed a hierarchical pipeline combining YOLOv11 segmentation, geometric rectification, and PaddleOCR for bedside monitors, reporting 99.08% field-level extraction accuracy on 6,498 images from Vietnamese ICUs [21]. Their pipeline, however, requires training two specialized detection models on thousands of annotated images, applies domain-specific post-processing heuristics, and evaluates only per-field accuracy without clinical metrics such as critical-error rate or Bland–Altman agreement. Chikhale and Mehendale (2025) linked vital-sign digitization to adaptive drug infusion using Tesseract OCR, reporting 99.87% accuracy on 3,000 synthetic monitor images for only two parameters (heart rate and blood pressure) [22]; no real-world clinical validation was performed. None of these studies has applied a vision–language model to the task.

Our study takes a different route: we treat monitor reading as a single-shot visual-information-extraction problem solved end-to-end by a general VLM, which requires no specialized detection model, no geometric rectification, and no domain-specific post-processing. Under this approach, a zero-shot frontier model reaches 0.997 exact match across all eight parameters simultaneously (a stricter metric than per-field accuracy), and a fine-tuned 9B open model achieves 0.994 on about 80 training images, two orders of magnitude fewer than the thousands required to train the detection stages of pipeline approaches such as that of Chau et al. (2025) [21]. The clinical evaluation (tolerance accuracy, critical-error rate, Bland–Altman agreement) we report fills a gap that none of the prior studies has addressed, and is essential for any system intended to feed values into clinical decision-making.

The frontier commercial models, accessible only through hosted APIs, achieve the highest raw accuracy in our study, but in the target setting deployability rather than accuracy is the binding constraint. Transmitting photographs of patient screens to a third-party cloud raises data-sovereignty and privacy obstacles that many institutions cannot clear [23]. Non-integrated operating rooms and wards may also lack the reliable connectivity a cloud API requires, and per-query pricing converts a continuous charting workload into a recurring operating cost. Once a 9B open model with a 0.46% adapter matches the best commercial model to within 0.3 points while running entirely on local hardware at near-zero marginal cost, the accuracy difference is clinically irrelevant and is outweighed by privacy, offline capability, cost, and controllability. That the strongest open model already matches the commercial models zero-shot is what makes this substitution feasible; LoRA closes the remaining gap without ceding control to a vendor.

Exact match is a stringent metric, but it treats a one-unit rounding error and a dangerous misread identically; the trustworthiness of the recipe therefore rests on the clinical analyses. Tolerance accuracy tracks exact match closely (0.959 ∼ 0.994), confirming that the fine-tuned model’s gains reflect correct reads rather than near-misses; its mean absolute error over the test parameters is effectively zero. The critical-error rate (the fraction of reads large enough to plausibly alter a clinical decision) falls from 0.0313 to 0.0063, with only two residual single-instance errors. Bland–Altman analysis localizes the remaining uncertainty to systolic blood pressure, specifically the invasive blood pressure (IBP)/non-invasive blood pressure (NIBP) selection error identified above rather than a digit-reading failure. The residual risk concerns which of two displayed pressures to report; it is addressable with a more explicit disambiguation prompt or a display-aware post-check. The digitization itself is clinically reliable, though IBP/NIBP source disambiguation remains a point where human confirmation is prudent.

The recipe’s resource footprint suggests that deployment need not require a server-class machine, though this should be read as a feasibility argument from measured resource consumption, not as a benchmarked claim. The adapted model consists of a 9B open backbone and a 43.3 M-parameter adapter. Fine-tuning (the heavier of the two phases) completed in roughly 26 minutes within a 28.5 GB memory budget on a single GPU; inference is lighter still. Because LoRA adds no inference-time latency once merged [6, 24], a quantized deployment of this model size [25] fits within the envelope of a single workstation-class or high-end consumer GPU, the kind of hardware a hospital can own and physically control.

Several limitations bound these conclusions. The study is single-center and modest in size (200 intraoperative images from one hospital, 40-image held-out test set). Although we use Wilson intervals and McNemar tests throughout, the absolute accuracies carry real sampling uncertainty, especially the one-instance separations among the top systems, and may not transfer to other sites, monitor vendors, or capture conditions. The device fleet is Mindray-dominated; performance on under-represented vendors is correspondingly less certain. The clinical metrics depend on the chosen tolerance and critical-error thresholds.

While we selected clinically motivated values, different thresholds would shift the absolute rates, though probably not the ranking. The commercial models are closed and accessed through evolving APIs, so their exact behavior is not perfectly reproducible; the version-specific open models we evaluate will also be superseded. Finally, the edge-deployment case is argued from resource footprint rather than measured on real hardware, and the recipe has not been evaluated in a live intraoperative documentation workflow.

Four directions follow from these limitations. External, multi-center validation spanning more monitor vendors and capture conditions would test generalization; such cohorts could potentially be trained without pooling raw data via privacy-preserving federation [23, 26]. A real edge benchmark should report latency, throughput, power, and quantization–accuracy trade-offs on embedded and consumer hardware. Integration into the hospital record through standard interfaces such as Health Level Seven/Fast Healthcare Interoperability Resources (HL7/FHIR) would allow digitized values to flow into the EHR and early-warning pipelines [5]. A prospective human-versus-model comparison measuring transcription error and time saved in routine practice would complement the static evaluation reported here.

## Code and Data Availability

The evaluation code, scoring module, fine-tuning configuration, and LoRA adapter weights are publicly available at https://github.com/gscfwid/AI_meet_monitor. The annotated dataset (monitor photographs and ground-truth labels) is available at https://huggingface.co/gscfwid/Qwen-3.5-9B-CareMonitor under MIT license.

## Acknowledgments

We thank the developers of the open-source models and frameworks whose work made this study possible: the Qwen team (Alibaba) for Qwen3.5-9B and Qwen3.6-27B, the MS-SWIFT team (ModelScope) for the fine-tuning framework, and the PaddlePaddle team (Baidu) for PaddleOCR. We also thank Google DeepMind for the Gemma open-weight models and Zhipu AI for GLM-5V-Turbo. Cloud GPU resources for fine-tuning were provided through Vast.ai. We thank Shan Gao, Jiaxin Peng and Xiaoxiang Chen for assistance with image acquisition. We thank the funding support provided by the Clinical Specialty Capacity Building Support Program at the First Affiliated Hospital of Sun Yat-sen University (R70026).

## Author Contributions

Gao S and He Q conceptualized the study and designed the evaluation framework. Wang P, Zhao X, and Yang B acquired the monitor photographs. Wang P and Zhang Z performed dual-reader ground-truth annotation, with discrepancies adjudicated by He Q. Gao S developed the evaluation pipeline, implemented the OCR baseline and VLM benchmarking code, conducted the LoRA fine-tuning experiments and ablation studies, performed the statistical analyses, and created all figures and visualizations. Gao S and Wang P wrote the original manuscript draft. Zhao X and He Q reviewed and modified the original manuscript. He Q and Feng X provided clinical oversight, supervised the study, and secured access to the operating-room environment and computational resources. All authors reviewed and edited the final manuscript and approved it for submission.

## Supplementary Tables

**Supplementary Table 1.** Per-parameter exact-match, tolerance, MAE, RMSE, critical-error rate, FN, FP for each model under basic/advanced prompts.

**Supplementary Table 2.** One-sided McNemar test for prompt effect (advanced ≥ basic) on 1,600 instances.

